# Early pregnancy in schools: a socio-ecological analysis of the determinants among teenage girls in Koudougou, Burkina Faso

**DOI:** 10.1101/2024.03.29.24305084

**Authors:** Wendkoaghenda Sophie Ramde, Patrice Ngangue, Tonye Kollo Appolinaire, Birama Apho Ly, Thieba Blandine Bonane

## Abstract

**Introduction:** Early pregnancy among adolescents in school has several consequences on the adolescent herself, her family, the whole community, and the child-to-be. This article explores the factors determining early pregnancy among 15-19-year-old secondary school students in Koudougou in Burkina Faso.

**Methods:** A descriptive and exploratory qualitative study was conducted through focus groups with adolescents and youth in schools and individual interviews with parents, teachers, health workers, and community leaders. The socio-ecological model guided all stages of the study. A thematic analysis of the recorded and transcribed data was conducted using Nvivo 12 software.

**Results:** A lack of knowledge and information and wrong perceptions about sexuality at the individual level; peer pressure and poverty at the interpersonal level; lack of awareness among teachers and students and health workers’ attitudes at the organizational level; the influence of new information technologies or the media and parents’ irresponsibility at the community level; and the insufficient of sexual and reproductive health services for adolescents as well as the lack of sanctions and law against early pregnancy at the political level were found as enablers. Barriers were the use of contraceptive methods and sexual abstinence; parents-children communication; teachers’ training on sexual sensitization, the creation of youth centers on school campuses and the introduction of sexual education courses; education through media and religion; willingness to introduce sexual education courses in school curricula and legal sanctioning of dealing and consuming drugs in schools.

**Conclusion:** The study highlighted that the problem of early pregnancy in schools can be solved by acting on the determinants at all levels of the socioecological model by implementing preventive strategies.

## Introduction

Adolescent pregnancy is a major social issue and a developmental problem because of its severe repercussions for the individual, their family and the community. It is a major concern for all of our societies, whether we are citizens of Northern or Southern states (1). Indeed, according to the World Health Organization (WHO), nearly 16 million adolescent girls worldwide, aged between 15 and 19 years, give birth to children each year, and more than 90% of adolescent pregnancies occur in developing countries (2). Early teenage pregnancy is also a significant problem in Africa. In sub-Saharan Africa, the prevalence of early pregnancy is 19.3% (3). Burkina Faso is not immune to early pregnancy among adolescents in general and, more specifically, in schools. Several policies, programs, and laws have been adopted to fight against early pregnancy in schools. Preventive actions have been implemented in Burkina Faso to address the problem. Despite these measures, the phenomenon remains a hot topic because it is rising. Although scientific literature is abundant on the issue of early pregnancy in schools and its various causes and consequences, the perceptions and factors that determine pregnancy in schools from adolescents’ point of view are poorly documented. Hence the interest in conducting the present study is to improve adolescents’ health status and reduce the risk of early pregnancy among school adolescents. According to a socioecological analysis, this observation justifies the realization of this study, which focuses on the determinants of early pregnancy in schools in Koudougou. Among all these lines of inquiry, our attention is focused on the following research question. What are the factors that determine the occurrence of early pregnancy among adolescents in schools in the city of Koudougou in Burkina Faso in 2021?

## Materials and Methods

The Socioecological Model (SEM) is a theoretical framework for understanding the multiple and interactive effects of personal and environmental factors that determine behaviors and identifying behavioral and organizational leverage points and intermediaries for health promotion within organizations. The combination of these factors influences an individual’s health status and explains the risk behaviors that the individual engages in (4). Facilitators and barriers to early pregnancy could operate at the intra-personal level (attitude, knowledge, adolescence, ability, beliefs and personality traits), at the interpersonal level (the groups to which we belong and the pressure these relationships exert on the way we behave, the family, social networks and friends that play a central role in the social development of individuals and explain their behaviours and health status,); at the organizational level (the school environment, organizations, schools, places of worship, public agencies and leisure groups etc.); at the community level (community, media, access to health services) and the political and public level (existing laws, policies and norms).

### Study design

This exploratory qualitative study explores the factors determining early pregnancy among adolescent girls in four secondary schools in Koudougou. The study was conducted from December 23, 2021, to February 4, 2022.

### Study population

The study population consisted of focus groups with adolescents in secondary schools and individual interviews with parents, teachers, health workers, and community leaders in the city of Koudougou, Burkina Faso. Two public secondary schools, the provincial and municipal secondary schools of Koudougou, one private secondary school, the Wendsongda private secondary school, and one religious secondary school, the Sainte Monique secondary school of Koudougou were choosen.

### Sampling

We used purposive sampling with maximum of variation.

### Recruitment Procedure

First, the investigator met with the school principals and the chief medical officer of the Koudougou health district to present the research project and to obtain their support for the study. During the meetings, the investigator notified each official of the targets (students and teachers, reproductive health officers and nurses) so that these different participants could be informed. At the level of community leaders, the investigator visited the Assemblies of God church, the Saint Augustine parish in Koudougou and the Dapoya mosque. The priest and the weekday pastor were approached, and a representative of the imam was met. The traditional chief was referred to the investigator by one of his nephews. To reach the parents of the students, the investigator contacted their representatives by telephone. The heads of the schools were previously notified to present the project, the modalities of their involvement and the ethical considerations associated with it, and to answer their questions and make an appointment for the interviews. The president of the Parent-Teacher Association had to choose the office members with their agreement. A health worker from each of the schools involved was interviewed. Of the participants who withdrew from the project, one had not obtained parental permission.

### Data Collection

The investigator used two data collection techniques for this study: individual interviews and focus groups. Informed consent respondents were interviewed using an interview guide by the principal investigator and a focus group guide by the principal investigator, who an assistant accompanied. The tools were validated and pretested.

### Data collection procedure

During the data collection process, we introduced ourselves to the participants. Then, we gave a brief presentation on the theme, the purpose, the general objective of the research and the modalities of the interview through the information note. Voluntarily, each participant completed and signed the free and informed consent form. Collecting information from the individual interviews and focus groups was done by smartphone recording the interviews. Observations with field notes were made using a logbook.

### Data processing and analysis

Data analysis was conducted using Nvivo 12 plus software and was based on the approach proposed by (5). who identified three major steps in qualitative data analysis. First, all interviews were transcribed verbatim on a Word file. Only one interview was transcribed from “Moor” into French. Some words were added during the transcription to indicate elements of the interview (laughter, silence, etc.). The transcription time was 30-45 minutes for each interview. The data were then coded. The coding was done by three (03) people, including the principal investigator and two students who were well-versed in the subject matter of the study. The concordance rate with merging the two databases of the principal investigator and the student was 98.77%. Finally, after the coding, the data were presented in a table format that facilitated the analysis work by generating an evolving and dynamic visual representation of the data reduction process.

### Scientific criteria

To avoid bias and ensure the quality of the results, the data were subjected to numerous interpretation tactics during the analysis and then to the verification of the conclusions as detailed in the scientificity criteria paragraph (5). These are the criteria of credibility, transferability, and internal consistency.

### Ethical considerations

Ethical approval was obtained from the Burkina Faso Health Research Ethics Committee (CERS) (N°2021-12-288). Authorization to conduct this study was obtained from the Centre-West Regional Health Directorate and the Provincial Directorate of Post-Primary and Secondary Education. Before enrolment, participants received all information related to the research and signed a consent form with the possibility to withdraw from the study without any disagreement.

## Results

We conducted six focus groups composed of 46 adolescents, i.e. three focus groups for girls and three focus groups for boys, each with 8 participants. The youngest participant was 14, and the oldest was 23 years old. Two teenage girls did not attend FGF1 and FGF3, respectively, without giving any reason, and 26 individual interviews were conducted. The average age of the adolescents was 17.80 years. The duration of the interviews varied between 14 and 45 minutes. The group discussions were longer and lasted between 62 and 115 minutes. The participants were distributed as follows.

The results of the qualitative data analyses enabled us to define two categories of factors that facilitate and hinder early pregnancy in schools. These factors operate at the individual, interpersonal, organizational, community and political levels.

### Facilitating factors for early pregnancy

By the socioecological model, the results reflect that the first level of explanation for the occurrence of pregnancy in schools is individual. Three (3) categories of factors were identified, namely knowledge and perception of teenage pregnancies, factors related to the level of information on sexual education and sexual and reproductive health (ignorance of sexuality concepts) and psychological and psychosocial factors referring to the teenager herself (the teenage crisis).

As a verbatim illustration of the factors linked to the level of information, a health worker said: “It is ignorance that makes many girls get pregnant because they think that there are periods when they cannot get pregnant, whereas they should protect themselves at all times” AS04.

At the interpersonal level, factors relating to the negative influence of peers and cultural factors come into play. This negative influence of peers is due to bad company, as stated by a parent (P10): “Bad company means that it is teenagers who have friends who may have more experience of sex and with whom the person is dating, which may lead them to want to follow in the footsteps of their friends.” Socio-cultural factors relate to parents’ relationship with their children about sexuality. The latter is still a taboo subject in many families. This means that children are exposed to unreliable sources and unwanted pregnancies. This is what the participants in focus group 03 acknowledged: “In my opinion, it’s also the lack of communication between young girls and their parents (…) there are other girls who are surprised to see their periods, so you don’t know that if you do it once, you can get pregnant, you’ll want to try your luck, and that’s the problem with people, but if the parents had taken the time to tell you that, you could have avoided pregnancies”.

In addition to sociocultural factors, there are socio-economic factors (poverty), as one adolescent girl explained: “In my opinion, it’s poverty because, at a certain age, parents don’t even take care to provide for your needs, to give you five hundred francs to pay for vania even, it’s a problem, so often you are obliged to do what you don’t want to do yourself to provide for your needs” FGF03.

At the organisational level, two factors are at work: teachers’/students’ attitudes and health professionals/ supervisors (lack of school awareness). Indeed, we are increasingly observing a disruption in the functioning of the school organisation, which is reflected in the guilty relationship between educators and learners and the non-involvement of health professionals as awareness-raising actors within schools. These words from an adolescent girl illustrate the inappropriate relationship between pupil and teacher: “The supervisors, you can come to school, and then there is no room, and the supervisor will tell you to give your number, and then he will give you a place, then you will call each other. So afterwards, accidents will happen” FGF02. Regarding the attitude of health professionals, the educational community in our study believes that their involvement in intra-school awareness-raising on pregnancy remains insufficient.

Two categories of factors are identified at the community level: the media’s negative influence and parents’ irresponsibility. Regarding the negative impact of the press, an adolescent girl stated (FG girl 01): “We can say that the telephone and television are the main causes of pregnancies because the pornographic sites that young people look at, such as telenovelas if they look at them, they can decide to try, and it is in trying that the pregnancy comes.” The irresponsibility of parents exposes adolescents to pregnancy because when they cannot provide for their children or are too absent to ensure their (sexual) education, the latter are vulnerable to certain proposals and do not have the correct information about sexuality. A nursery health worker said: “We have noticed that we parents are not too involved, especially in the sexual education of children, whether they are girls or boys. It’s a taboo between parents and children. So, when children don’t have information just from their parents, they will get information from their friends, which can also expose the girl” (AS03).

At the political level, we have an insufficient supply of sexual and reproductive health services, as evidenced by the words of AS04 “There is an insufficiency of sexual health services for young people everywhere (…)” and the absence of laws. Community leaders believe that the lack of sanctions against victims and perpetrators of underage pregnancies encourages the occurrence of pregnancies among this group. This suggests that early pregnancy in schools is not yet genuinely perceived as a public health problem in Burkina Faso.

### Factors hindering the occurrence of early pregnancy in schools

The factors that hinder early pregnancies in schools, or barrier factors, as well as facilitating factors, operate at the same levels.

At the individual level, we have two categories of factors: the use of contraceptive methods and the practice of sexual abstinence. Most adolescents approve of this because awareness is insufficient. This is illustrated by the words of a pupil from a focus group: “I think that to solve these factors, it is the contraceptive methods that will be easier to help young girls (…)” (FGF03). Sexual abstinence is mentioned by health workers, teachers and religious people as a way not only to avoid early pregnancies but also to keep girls in school.

At the interpersonal level, parental influence was identified as a factor in protecting adolescent girls from early pregnancy. This sex education concerns both young boys and girls. To do so, parents must be available and open to dialogue. Parents must also control their children’s exposure to the media. They should also be helped by the state, which should censor and block inappropriate programs. One teacher said in this regard, “(…) because elsewhere, I see, you can’t access specific sites, it doesn’t fit with their value in some countries, if we can censor certain things too, I think it can help. (…)”. (Ens01).

At the organisational level, a category of factors related to the attitude and skills of health professionals and school supervisors was identified. In fact, at the school level, the participants welcomed the introduction of sex education courses and the enrichment of the curricula of certain subjects because “It allows young people to get to know each other, and the girl to understand how her organs work” (LC04). It would also be necessary, according to the participants, to ensure that the capacities of school actors in the area of sexuality are strengthened so that they can take on the task of raising awareness about pregnancy in schools. In the same vein, the creation of centres for young people within schools will enable health professionals to have more impact in their actions because these centres “(…) accompany with contraceptive methods, condoms, depo-provera” (AS01).

At the community level, participants noted the influence of the media and religion as protective factors. Indeed, the media are useful in raising awareness of a large public against early pregnancies. In this regard, a health worker stated, “The media can help us with the various awareness campaigns; last year, there were at least three to four radio stations in sync during the family planning campaign (AS 01). As for religion, its precepts and commandments, when followed to the letter, can protect young girls from early pregnancy. For example, as this adolescent girl states, “In the church, in the Bible, you are told not to practice adultery; that the girl must remain a virgin until she is married, and all this can also help in any case if the girl listens and wants to put it into practice (…).” (FGF01)

At the policy level, the participants cited two categories of factors: the introduction of a sex education module in the school curriculum and the fight against the use of drugs and alcohol in schools. While regretting the inconsistency and absence of sex education modules in school curricula and programmes, the participants, especially the teachers, are convinced that the introduction of a sex education course would help to reduce the rate of early pregnancies in schools. Parents, school staff and students believe that introducing and using drugs and alcohol in schools should be vigorously opposed, especially during cultural weeks. They recommend introducing legislation to deal with offenders in this regard.

## Discussion

Early school pregnancies are a real sexual and reproductive health problem in Burkina Faso. Analysis of the determinants of these pregnancies has enabled us to make findings at various levels. These findings are corroborated by previous research.

The factors that facilitate early pregnancy in schools are of several kinds and can be observed at different levels. At the individual level, adolescent students’ insufficient or erroneous knowledge of sexuality-related issues appears to be the primary factor explaining school pregnancy. This result is similar to those of (6,7) and (8). This situation could be explained by the lack or inadequacy of school awareness and the persistence of prejudices about late sexuality. Also, the fact that sex is still a taboo subject (9) mentions the non-enrolment of some parents and the non-attendance of parents for their children due to the demands of the capitalist society. Also, according to the study by (10), 38 students (47%) of the interviewees, confessed to having contracted their pregnancy out of ignorance.

Psychological and psychosocial factors show that adolescence is a determinant of early school pregnancy. Indeed, during this period adolescents undergo physiological, psychological and emotional changes that are characterised by curiosity, neglect and a high level of sexual pleasure seeking. Our results are comparable to those of (8,11–13). At the interpersonal level, the factors related to the adolescents’ entourage make us understand the negative influence of peers on the occurrence of early pregnancies in schools. Several studies similar to our research agree with this (6,8,10,14,15).

Socio-cultural factors include poor parent-child communication about sexuality. There are several reasons for this: The first is that sex is still a taboo subject that should not be discussed with children at the risk of indirectly encouraging them to engage in early sexuality. The second reason is the parents’ lack of mastery of the subject due to their under-schooling and the conservative education they received from their parents. The primacy of professional occupations over (sexual) education means that adolescent children are left to their own devices in matters of sexuality and consequently find themselves taking advice from peers who are not better informed and wise, thus giving free rein to their impulses. This result is similar to that of (6,8,14,16–18).

The socio-economic status of the family, particularly poverty, is a risk factor for early pregnancy in schools. Indeed, when school-related needs that poor parents cannot meet, adolescent girls engage in risky sexual behaviour. Studies in Burkina Faso, Cameroon,

Guinea, Senegal, Burundi and Côte d’Ivoire have also shown that poverty is a significant risk factor for early pregnancy in schools (6,8,10,11,14,16,19).

At the organisational level, our result about attitudes/skills of health professionals/ supervisors shows insufficient awareness in schools. This situation is due to the lack of funding for awareness-raising activities at the commune level to cover all the schools, the lack of nurseries and the absence of programmes and specific strategies for preventing and managing early pregnancies. This result is similar to that of a study conducted in Burundi by (8).

The results about teacher/student attitudes show inappropriate teacher/student relationships. Teachers no longer play their role as educators. Indeed, instead of raising awareness and putting adolescent girls on track, they are responsible for their defection by being guilty of reprehensible relationships. They use their position of authority to harass or abuse the vulnerability of adolescent girls.

They also often give in to the advances of students. It would therefore be advisable for the school administration to protect pupils from guilty relationships with teachers, to encourage and pay attention to any form of harassment, violence or abuse in the school environment by sanctioning guilty teachers and bringing the matter before the teachers’ disciplinary council.

Studies conducted in Burundi, Burkina Faso, have shown, as our study has, that closeness between teachers and pupils is a risk factor for early pregnancy (6,8,20).

At the community level, our findings related to the influence of media, mobile phones, and movies show that these expose adolescents to risky sexual behaviours that can lead to early pregnancy in schools. Through these media, they learn at an early age about deviant sexuality, which they will be tempted to practice. The telephone, for example, opens the door to meetings and exchanges with a purely sexual connotation. Studies in Burundi and Burkina Faso agree with our finding that the media exposes adolescents to adult sexual behaviour (6,8).

Our findings concerning parental influence show the absence of sex education in the family. This is because although families continue to educate and socialise children, sex education remains the poor relation of parental educational provision. Our result is similar to that of a study in Burundi, which shows that the values advocated by parents can play an important role in preventing pregnancies and being an enabling factor (8).

At the policy level, our findings concerning the availability of ASRH/FP services show the inadequacy of Adolescent and Youth Sexual and Reproductive Health/Family Planning (ASRH/FP) services. There are several reasons adolescents do not use health services: fear of being discovered by parents, lack of privacy, doubts about respecting confidentiality, and poor reception by service providers. We can also add the approximate handling by the providers and the fear of being judged by the providers for their choices and actions. In addition to these causes, there are prejudices about the target audience of ASRH/FP services, lack of information about the location and services offered, fear of medical examinations, the distance between adolescents’ homes and the clinics, restrictions in terms of policies and laws relating to young people and contraception, and the high cost of services, etc.

Similar studies in Benin, Senegal and Central Africa have shown that young people do not have sufficient access to reproductive health services (11,14,18).

Our findings related to the legal and penal framework attest to the impunity of perpetrators and victims of pregnancy. The poverty and misery characterizing the populations could explain this’ tolerance.’ This legal vacuum was highlighted in the study conducted in Burundi (8).

Other types of factors are linked to the occurrence of early pregnancies. These are barriers, hindering or protective factors. They can contribute to limiting early pregnancy in schools if the various actors and stakeholders focus on them.

At the individual level, our results concerning contraceptive use attest to adolescents’ knowledge and use of contraceptive methods. This result is similar to that of (6,8). This could be explained by contraceptive methods allowing girls to better manage their sexuality by taking the necessary precautions to avoid early pregnancies, Sexually Transmitted Infections and Diseases. In addition to using contraceptive methods at the individual level, sexual abstinence is another protective factor against pregnancy in the school environment. This practice allows the young girl to complete her schooling and avoid early pregnancy and STIs. At the interpersonal level, the primary protective factor is the influence of parents. Indeed, when parents educate their children (at an early age) about sexuality, control their exposure to the media and screen their dating, children tend to develop less risky sexual behaviour. Similar results were found by (21).

At the organisational level, we note that the introduction of sex education courses in schools helps to foster attitudes of individual and collective responsibility. Our results are in line with those of (6,10,13,19). Our results also point to the establishment of a youth centre within schools. These centres could serve as a relay for awareness raising/care and fill some of the unmet needs of these young people, as noted by (6).

At the community level, the media stand out as a protective factor against school pregnancy because of their information, training and awareness-raising roles (6,8,13). This is hardly surprising since the media remains an unavoidable channel for disseminating information. Religion is also an important factor at the community level; indeed, all religions advocate moral and social values and urge unmarried youth and adolescents to abstain from sex. But also, religion recommends that children respect and follow their parents’ advice and that the latter assume responsibility for their children’s education. This result is superimposed on the study by (6).

At the policy level, our results and those of (6) confirm that introducing sex education in school curricula and programs will enable students to develop specific skills and knowledge to make responsible decisions about sexuality.

The occurrence of pregnancy in schools involves actors at different levels. This implies that the school administration sensitises students on adolescent pregnancy through discussion forums, after-school and extra-curricular activities, equips the nurseries and involves their managers and external actors in sensitisation. As for the parents, they must detach themselves from sex and its education. They must also provide for their children and teach them the culture of reporting harassment and (sexual) abuse. In the case of community leaders, they should advocate for the punishment of perpetrators of teenage pregnancy, especially in schools. Finally, adolescents must practice abstinence or, failing that, protect themselves during sexual relations. In the event of rape, they must denounce the perpetrators.

Our study has some limitations. Firstly, as qualitative research, the results of this study are not generalizable, as only four of the 176 institutions in the province were included in the study. Although the individual interviews and focus groups aimed at saturating the information, likely, some aspects of interest were not addressed. There was a difference in depth, richness and length between the individual interviews and the focus groups, which suggests that the focus group dynamics were more effective in eliciting discourse, perceptions and explicit views on the subject than the in-depth individual interviews.

Also, our subjectivity could taint the data collection, which is essentially based on the perceptions of the interviewed participants and the subjective interpretation of the research results. In addition, the answers given by the participants may not be objective; sexuality is still taboo; therefore, they may feel embarrassed, which may induce declarative bias or social conformity bias.

## Conclusion

This research has shown that the consequences of pregnancy in schools are academic, social, economic, physical and mental health. Early pregnancies result from several facilitating factors such as ignorance, adolescence at the individual level, poverty, poor company, lack of communication with parents at the interpersonal level, insufficient awareness at the organizational level, the media at the community level, and the political level, the inadequate availability of Adolescent and Youth Sexual and Reproductive Health/Family Planning services. As factors that hinder the occurrence of pregnancies in schools, our study highlighted sexual abstinence, the use of contraceptive methods at the individual level, parent/child communication at the interpersonal level, sex education courses in schools at the organizational level, and the availability of ASRH/FP services for adolescents at the policy level. Therefore, there is a need to develop programs to prevent early pregnancy among adolescents from the individual to the policy level. In addition, it is also essential to improve the provision of adolescent-friendly reproductive health services and greater collaboration between schools and medical facilities. Also, each family must take the dialogue to heart to give adolescents the right information for responsible sexual behavior. Government, parents, school authorities, and health professionals must all work together to break the taboos and dare to talk to young people about sex education and contraceptives.

## Data Availability

All data produced in the present work are contained in the manuscript

## List of acronyms and abbreviations

ICTs: New Information Technologies
SEM: Socio-Ecological Model
PTA: Parent Teacher Association
CERS: Health Research Ethics Committee
ASRH/FP: Adolescent and Youth Sexual and Reproductive Health/Family Planning

---

**Table 1.**
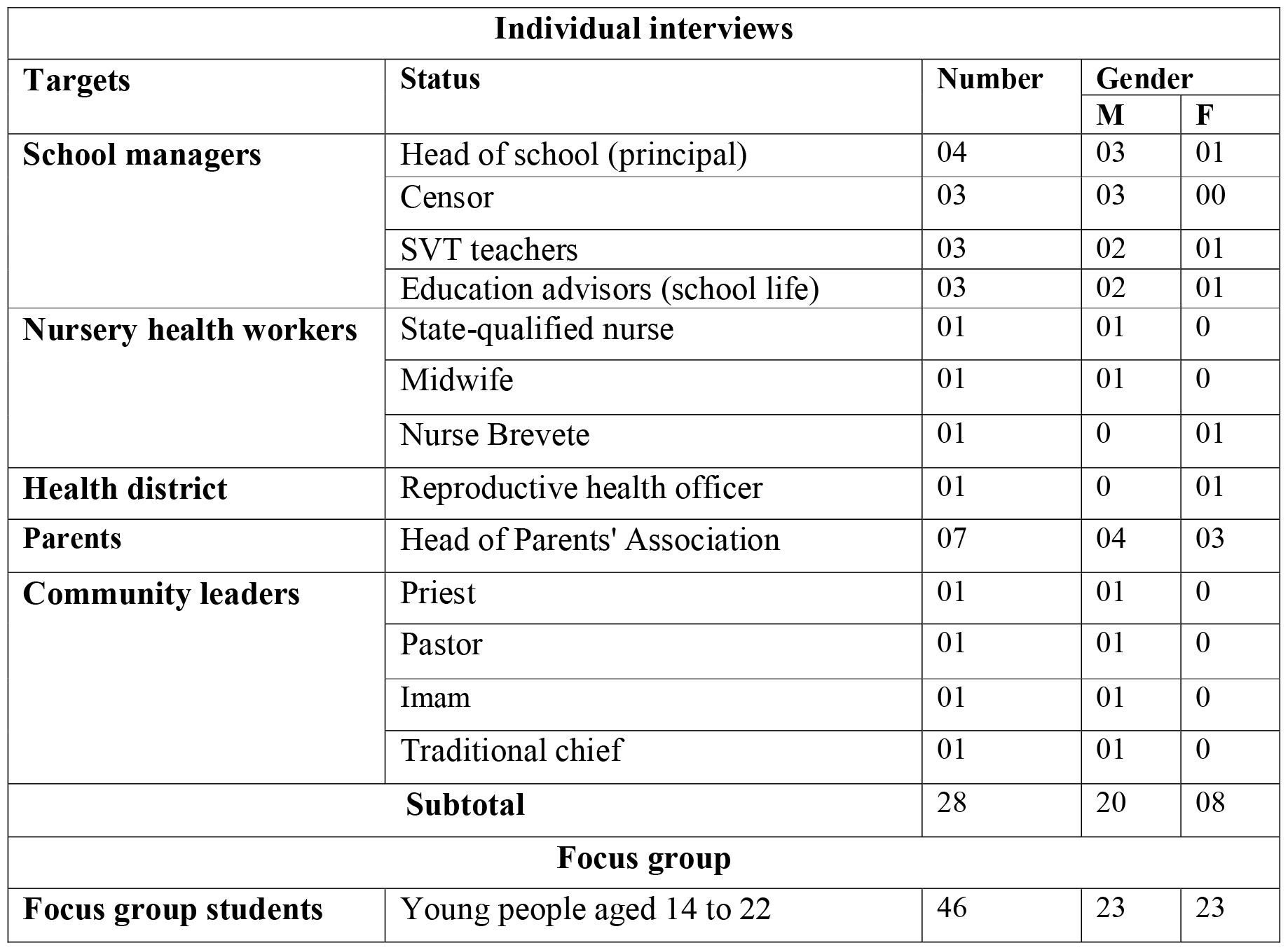
Distribution of interviewees.

| **Individual interviews** |  |  |  |  |
| --- | --- | --- | --- | --- |
| **Targets** | **Status** | **Number** | **Gender** |  |
|  |  |  | **M** | **F** |
| **School managers** | Head of school (principal) | 04 | 03 | 01 |
|  | Censor | 03 | 03 | 00 |
|  | SVT teachers | 03 | 02 | 01 |
|  | Education advisors (school life) | 03 | 02 | 01 |
| **Nursery health workers** | State-qualified nurse | 01 | 01 | 0 |
|  | Midwife | 01 | 01 | 0 |
|  | Nurse Brevete | 01 | 0 | 01 |
| **Health district** | Reproductive health officer | 01 | 0 | 01 |
| **Parents** | Head of Parents' Association | 07 | 04 | 03 |
| **Community leaders** | Priest | 01 | 01 | 0 |
|  | Pastor | 01 | 01 | 0 |
|  | Imam | 01 | 01 | 0 |
|  | Traditional chief | 01 | 01 | 0 |
| **Subtotal** |  | 28 | 20 | 08 |
| **Focus group** |  |  |  |  |
| **Focus group students** | Young people aged 14 to 22 | 46 | 23 | 23 |

